# SARS-CoV-2 Viremia is Associated with Distinct Proteomic Pathways and Predicts COVID-19 Outcomes

**DOI:** 10.1101/2021.02.24.21252357

**Authors:** Yijia Li, Alexis M. Schneider, Arnav Mehta, Moshe Sade-Feldman, Kyle R. Kays, Matteo Gentili, Nicole C. Charland, Anna L.K. Gonye, Irena Gushterova, Hargun K. Khanna, Thomas J. LaSalle, Kendall M. Lavin-Parsons, Brendan M. Lilly, Carl L. Lodenstein, Kasidet Manakongtreecheep, Justin D. Margolin, Brenna N. McKaig, Blair A. Parry, Maricarmen Rojas-Lopez, Brian C. Russo, Nihaarika Sharma, Jessica Tantivit, Molly F. Thomas, James Regan, James P. Flynn, Alexandra-Chloé Villani, Nir Hacohen, Marcia B. Goldberg, Michael R. Filbin, Jonathan Z. Li

## Abstract

**Background:** Severe Acute Respiratory Syndrome Coronavirus 2 (SARS-CoV-2) plasma viremia has been associated with severe disease and death in coronavirus disease 2019 (COVID-19) in small-scale cohort studies. The mechanisms behind this association remain elusive.

**Methods:** We evaluated the relationship between SARS-CoV-2 viremia, disease outcome, inflammatory and proteomic profiles in a cohort of COVID-19 emergency department participants. SARS-CoV-2 viral load was measured using qRT-PCR based platform. Proteomic data were generated with Proximity Extension Assay (PEA) using the Olink platform.

**Results:** Three hundred participants with nucleic acid test-confirmed COVID-19 were included in this study. Levels of plasma SARS-CoV-2 viremia at the time of presentation predicted adverse disease outcomes, with an adjusted odds ratio (aOR) of 10.6 (95% confidence interval [CI] 4.4, 25.5, P<0.001) for severe disease (mechanical ventilation and/or 28-day mortality) and aOR of 3.9 (95%CI 1.5, 10.1, P=0.006) for 28-day mortality. Proteomic analyses revealed prominent proteomic pathways associated with SARS-CoV-2 viremia, including upregulation of SARS-CoV-2 entry factors (ACE2, CTSL, FURIN), heightened markers of tissue damage to the lungs, gastrointestinal tract, endothelium/vasculature and alterations in coagulation pathways.

**Conclusions:** These results highlight the cascade of vascular and tissue damage associated with SARS-CoV-2 plasma viremia that underlies its ability to predict COVID-19 disease outcomes.

## Introduction

With coronavirus disease-2019 (COVID-19) causing over two million deaths globally by early 2021^1^, there remains an urgent need to elucidate disease pathogenesis to improve clinical management and treatment. There is increasing evidence that COVID-19, caused by the severe acute respiratory syndrome coronavirus 2 (SARS-CoV-2) virus, frequently manifests pathology beyond the pulmonary tract ^2-4^. In both immunocompromised and immunocompetent hosts, SARS-CoV-2 nucleic acids have been detected across a broad range of extrapulmonary sites, including spleen, heart, liver, and intestinal tract ^5-9^. In addition, endothelial cells are known to express ACE-2 and some reports have suggested that direct infection of endothelial cells may be leading to a hypercoagulable state with vascular and downstream organ damage. Furthermore, viremia has been implicated in transplacental transmission ^7,10^. These reports suggest that dissemination of infection outside of the respiratory tract into the circulatory system may be a critical step for COVID-19 pathogenesis.

We and others have previously demonstrated that SARS-CoV-2 plasma viremia in hospitalized patients is associated with severe disease and death ^11-14^. However, these studies have been limited by sampling late during the disease course and relatively small sample sizes. Here, we performed plasma SARS-CoV-2 viral load quantification, proteomic analysis, and assessed neutralizing antibody titers in a large cohort of emergency department (ED) patients enrolled at the time of initial presentation. We evaluated whether levels of SARS-CoV-2 viremia could predict COVID-19 disease outcomes after adjusting for multiple potential confounders. We also performed proteomic analysis to reveal prominent pathways that are upregulated in the setting of plasma viremia and determined the relationship between plasma SARS-CoV-2 viral load and levels of neutralizing antibodies.

## Results

### Baseline participants characteristics

This cohort consisted of 306 participants with a molecular diagnosis of COVID-19, of which 300 participants had successful plasma SARS-CoV-2 viral load quantification and thus were included in this current analysis. Baseline characteristics were reported in our prior study ^15^ and summarized in Table 1. Thirty-nine percent of participants were 65 years or older and about half of participants were female. Eleven percent of participants had morbid obesity (body mass index [BMI] ≥40 kg/m^2^), 47% had a diagnosis of hypertension and 36% with diabetes. Fifty-three out of 300 participants (18%, Figure 1A) had a baseline SARS-CoV-2 viral load above the limit of quantification (2 log_10_ copies/ml). Individuals with quantifiable SARS-CoV-2 viral load at the time of ED presentation were of older age, had higher rates of diabetes, and had clinical laboratory values consistent with higher disease severity, including lower lymphocyte count, and higher creatinine, C-reactive protein (CRP), and troponin (Table 1). Median time between symptom onset and ED presentation was 7 days (interquartile range [IQR], 4, 11) and comparable between individuals with viral load above and below the limit of quantification (Figure 1B and Supplementary Figure S1). Quantified SARS-CoV-2 viral load at the time of ED presentation was correlated with older age, lower lymphocyte count, higher inflammatory markers including CRP, D dimer, Lactate dehydrogenase (LDH), and with both renal and liver dysfunction (Figure 1C).

**Table 1.** Summary of baseline characteristics

|  | Total<br>(n=300) | Viremic within<br>quantification range<sup>a</sup> (n=53) | Aviremic or below<br>quantification range (n=247) | P |
| --- | --- | --- | --- | --- |
| Age (n, %) |  |  |  | **0.04** |
| <50 years | 96 (32.0) | 10 (18.9) | 86 (34.8) |  |
| 50-64 years | 86 (28.7) | 15 (28.3) | 71 (28.7) |  |
| ≥65 years | 118 (39.3) | 28 (52.8) | 90 (34.4) |  |
| Female (n, %) | 144 (48.0) | 20 (37.7) | 124 (50.2) | 0.10 |
| Non-Caucasian (n, %) | 150 (50.0) | 31 (58.5) | 119 (48.2) | 0.17 |
| Morbid obesity<sup>b</sup> (BMI≥40<br>kg/m<sup>2</sup>, n, %) | 33 (11.8) | 7 (13.7) | 26 (11.4) | 0.19 |
| Heart diseases (n, %) | 46 (15.3) | 6 (11.3) | 40 (16.2) | 0.37 |
| Lung diseases (n, %) | 64 (21.3) | 6 (11.3) | 58 (23.5) | **0.05** |
| Hypertension (n, %) | 143 (47.7) | 28 (52.8) | 115 (46.6) | 0.41 |
| Diabetes (n, %) | 108 (36.0) | 28 (52.8) | 80 (32.4) | **0.005** |
| Immunocompromised<br>condition (n, %) | 25 (8.3) | 5 (9.4) | 20 (8.10) | 0.75 |
| Lymphopenia <1000<br>cells/mm<sup>3</sup> (n, %) | 149 (49.7) | 33 (62.3) | 116 (47.0) | **0.04** |
| Creatinine elevation >1.20<br>mg/dl (n, %) | 64 (21.3) | 17 (32.1) | 47 (19.0) | **0.04** |
| CRP<sup>c</sup>>100 mg/dl | 146 (50.7) | 38 (73.1) | 108 (45.8) | **<0.001** |
| D-dimer<sup>d</sup>>1000 ng/ml (n, %) | 151 (53.2) | 34 (66.7) | 117 (50.2) | **0.03** |
| Troponin elevation within 72<br>hours (n, %) | 24 (8.0) | 8 (15.1) | 16 (6.5) | **0.04** |
| Baseline SARS-CoV-2 viral<br>load, log copies/ml (median,<br>IQR) | N/A | 2.68 (2.39, 3.63) | N/A | N/A |
| Percentage of detectable but<br>not quantifiable viremia (n, %) | 53 (17.7) | 0 (0.0) | 53 (21.5) | N/A |

a, quantification range for viremia is $\geq 2.0$ log copies/ml.
b, 280 participants with available BMI.
c, 288 participants with available CRP.
d, 284 participants with available D-dimer.
BMI, body mass index; CRP, C reactive protein; SARS-CoV-2, severe acute respiratory syndrome coronavirus 2; IQR, interquartile range; N/A, not applicable.

**Figure 1.**
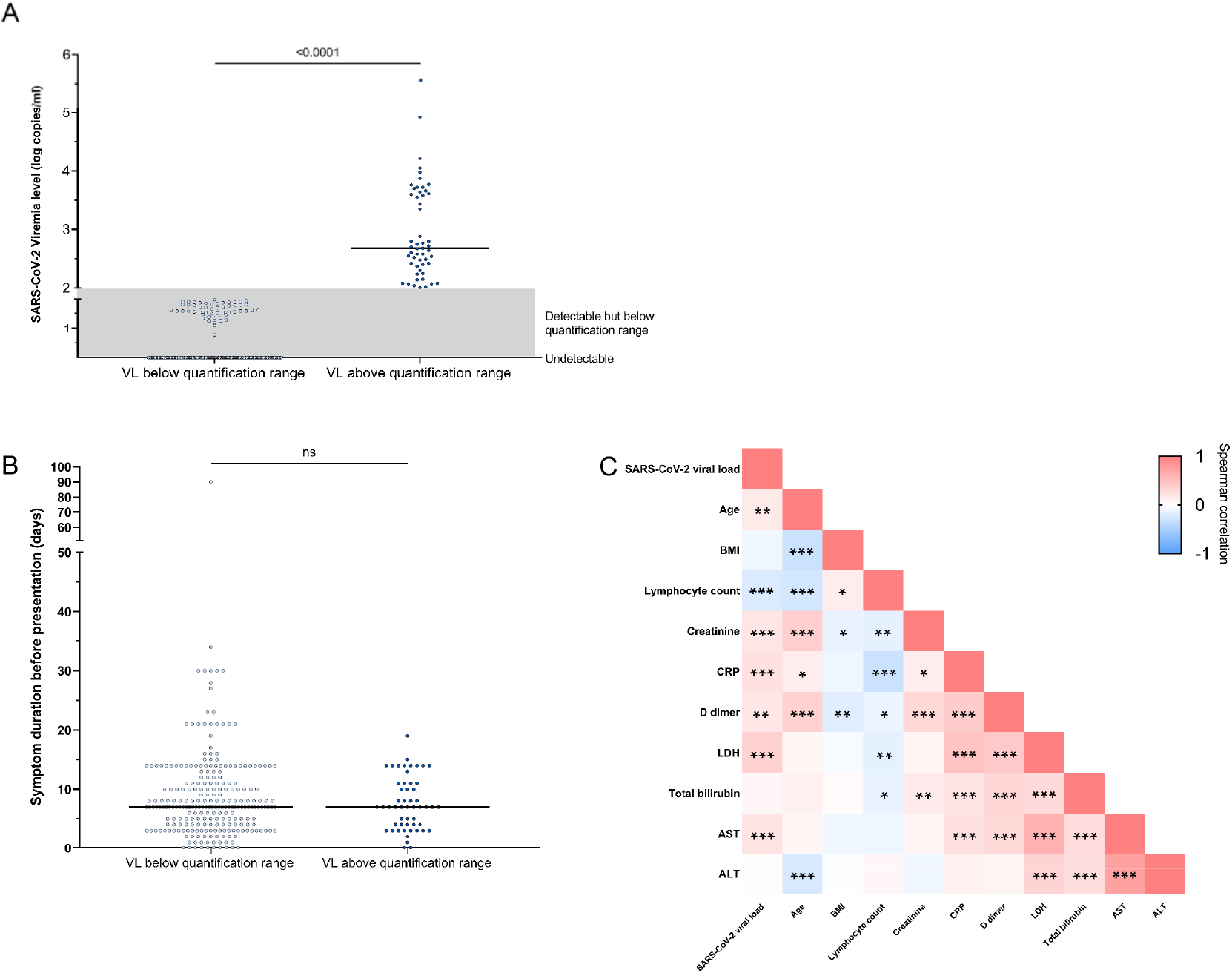
SARS-CoV-2 viremia at Day 0. (A) Distribution of SARS-CoV-2 viral load. 53 participants had viremia within the quantification range with median viral load 2.68 log copies/ml; 247 participants had viral loads (VLs) below the range of quantification or detection. (B) Duration between symptom onset and ED presentation was comparable between the viremic (quantifiable) and the aviremic/viremic (unquantifiable) group. (C) Pairwise correlation heatmap between viral load and baseline factors (Spearman’s rank correlation coefficient). *, P<0.05; **, P<0.01; ***, P<0.001.

### SARS-CoV-2 viremia at the time of ED presentation predicted adverse clinical outcomes during the hospitalization

Elevated SARS-CoV-2 viremia ≥2 log_10_ copies/ml at the time of ED presentation was a strong predictor of maximal COVID-19 disease acuity within 28 days of enrollment. Those with elevated viral load were significantly more likely to have severe disease (82% vs. 26%, P<0.001, Figure 2A), which included those who died or required invasive mechanical ventilation. Participants with SARS-CoV-2 viral loads <2 log_10_ copies/mL were further categorized into those with detectable viral load below the limit of quantification or with undetectable viral load (aviremic). This revealed a dose-dependent effect of viremia on adverse outcomes (Figure 2B). Higher levels of SARS-CoV-2 viremia upon ED presentation were associated with increased severity at all timepoints measured - days 0, 3, 7, and 28 (Supplementary Figure S2). 28-day mortality was 32% in the high viral load group and 9.7% in the low viral load group (P<0.001). Higher plasma viral load was also consistently associated with higher risk of severe disease and death across age groups (Supplementary Figure S3).

**Figure 2.**
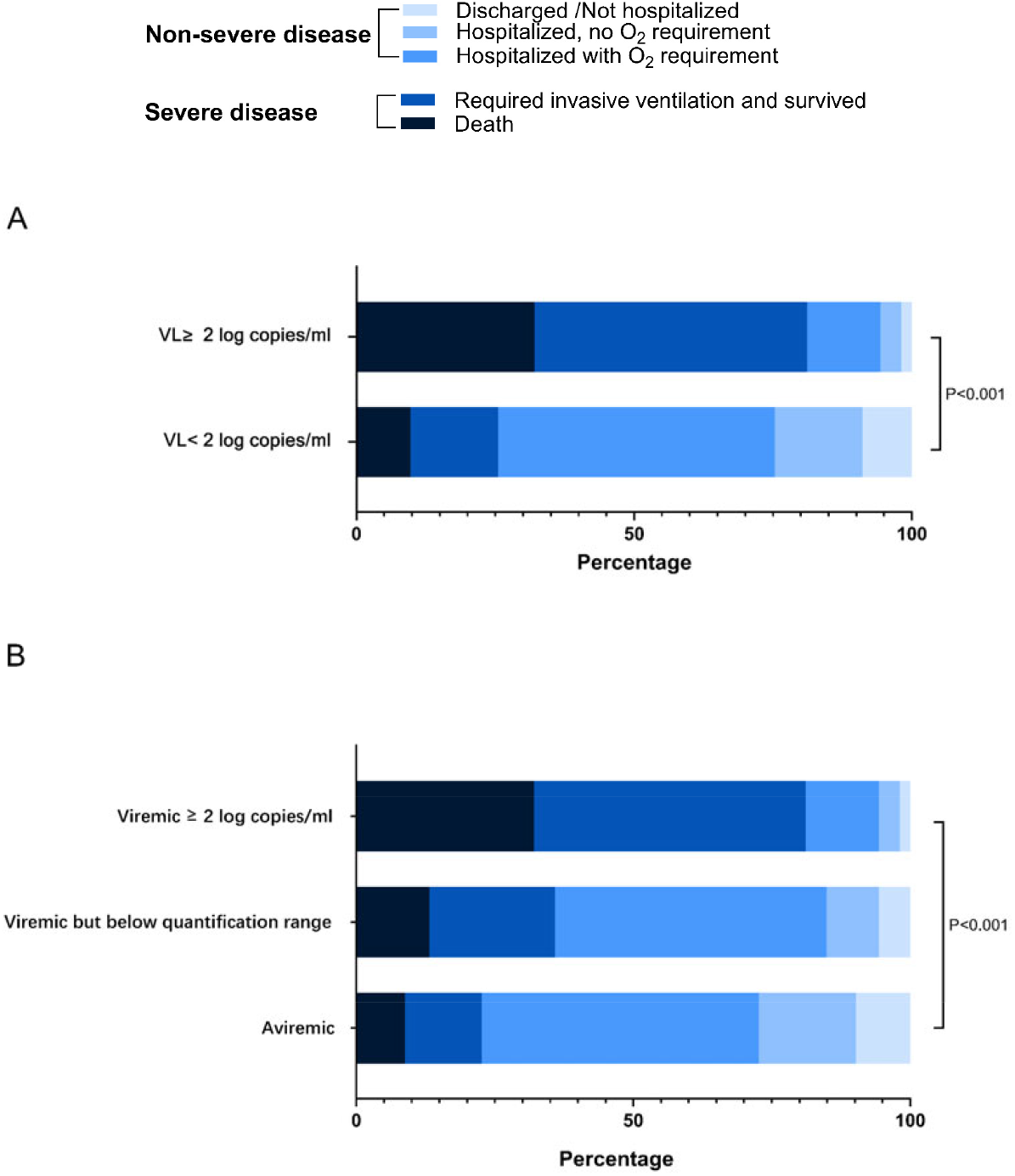
Association between baseline SARS-CoV-2 viral load and maximal disease severity (Acuity_max_). (A) Disease severity categorized by viral load above and below the quantification limit (≥ 2 log_10_ copies/ml vs. < 2 log_10_ copies/ml). (B) Disease severity categorized by viral load within the quantification range, below the quantification range but detectable, or aviremic.

We also assessed the impact of SARS-CoV-2 by univariate and multiple logistic regression for severe disease. Viremia ≥2 log_10_ copies/ml had an OR of 12.6 (95% CI 6.0, 26.5, P<0.001) in univariate logistic regression for severe disease (Table 2). After adjusting for other baseline variables with a P value <0.1 in univariate analyses, viremia remained significantly associated with severe disease, with an adjusted OR (aOR) of 10.6 (95% CI 4.4, 25.5, P<0.001). Similarly, viremia ≥2 log_10_ copies/ml was strongly associated with death within 28 days (Table 2), with an aOR of 3.9 (95% CI 1.5, 10.1, P=0.006) in multivariate analysis. The results were consistent when viral load was categorized into 3 strata (2 log_10_, detectable below 2 log_10_ and aviremic) and when analyzed as a continuous variable (Supplementary Table S1). Each log_10_ increase in viral load was associated with an aOR 2.49 of severe disease (P<0.001) and aOR 1.46 of death (P=0.01). Finally, higher viral load was also associated with higher risk of death at day 28 by Cox proportional hazard modelling (adjusted hazard ratio [aHR] 4.0, 95% CI 1.9, 8.7, P<0.001, Supplementary Figure S4). We performed logistic regression to evaluate demographic and laboratory variables associated with SARS-CoV-2 viremia. In multivariate analysis, only diabetes and CRP>100mg/dl were associated with viremia (Supplementary Table S2).

**Table 2.** Factors associated with severe COVID-19 and death.

|  | Univariate OR (95% CI) | P | Multivariate OR (95% CI) | P |
| --- | --- | --- | --- | --- |
| **Severe disease** |  |  |  |  |
| SARS-CoV-2 viremia |  |  |  |  |
| Aviremic or below quantification range | Reference |  | Reference |  |
| Viremic $\geq 2$ log copies/ml | 12.56 (5.96, 26.46) | <0.001 | 10.59 (4.40, 25.51) | **<0.001** |
| Age |  |  |  |  |
| <50 years | Reference |  | Reference |  |
| 50-64 years | 2.01 (1.00, 4.04) | 0.049 | 1.06 (0.43, 2.59) | 0.91 |
| $\geq 65$ years | 5.32 (2.82, 10.06) | <0.001 | 2.58 (1.02, 6.52) | 0.045 |
| Female (male as reference) | 0.71 (0.44, 1.14) | 0.16 |  |  |
| People of color (Caucasian as reference) | 1.42 (0.88, 2.29) | 0.15 |  |  |
| Morbid obesity (BMI$\geq 40$ kg/m<sup>2</sup>) | | | | |
| No | Reference |  |  |  |
| Yes | 0.86 (0.40, 1.85) | 0.69 |  |  |
| Unknown | 0.43 (0.14, 1.32) | 0.14 |  |  |
| Heart diseases | 1.86 (0.98, 3.50) | 0.06 | 2.05 (0.79, 5.29) | 0.14 |
| Lung diseases | 0.49 (0.26, 0.92) | 0.03 | 0.32 (0.13, 0.80) | **0.02** |
| Hypertension | 2.09 (1.29, 3.38) | 0.003 | 0.87 (0.41, 1.82) | 0.70 |
| Diabetes | 1.97 (1.21, 3.21) | 0.007 | 1.58 (0.80, 3.11) | 0.19 |
| Immunocompromised conditions | 2.53 (1.11, 5.80) | 0.03 | 1.93 (0.69, 5.41) | 0.21 |
| Lymphopenia <1000 cells/mm<sup>3</sup> | 2.83 (1.73, 4.64) | <0.001 | 2.00 (1.04, 3.84) | **0.03** |
| Creatinine elevation >1.20 mg/dl | 3.93 (2.21, 7.00) | <0.001 | 2.12 (0.92, 4.90) | 0.08 |
| CRP>100 mg/dl |  |  |  |  |
| No | Reference |  | Reference |  |
| Yes | 4.75 (2.80, 8.08) | <0.001 | 3.15 (1.60, 6.19) | **0.001** |
| Unknown | 0.85 (0.18, 4.11) | 0.84 | 1.72 (0.06, 46.51) | 0.75 |
| D-dimer>1000 ng/ml |  |  |  |  |
| No | Reference |  | Reference |  |
| Yes | 3.21 (1.92, 5.39) | <0.001 | 1.40 (0.71, 2.76) | 0.34 |
| Unknown | 0.79 (0.21, 2.96) | 0.73 | 0.63 (0.03, 12.03) | 0.76 |
| Troponin elevation within 72 hours | 6.41 (2.46, 16.70) | <0.001 | 3.74 (1.01, 13.83) | **0.048** |
| **Death within Day 28** |  |  |  |  |
| SARS-CoV-2 viremia |  |  |  |  |
| Aviremic or below quantification range | Reference |  | Reference |  |
| Viremic $\geq 2$ log copies/ml | 4.39 (2.15, 8.96) | <0.001 | 3.86 (1.47, 10.14) | **0.006** |
| Age |  |  |  |  |
| <50 years | Reference |  | Reference |  |
| 50-64 years | 3.43 (0.35, 33.65) | 0.29 | 1.98 (0.16, 25.25) | 0.60 |
| $\geq 65$ years | 43.39 (5.82, 323.31) | <0.001 | 22.61 (2.07, 246.40) | **0.01** |
| Female (male as reference) | 0.83 (0.43, 1.60) | 0.57 |  |  |
| People of color (Caucasian as reference) | 0.47 (0.24, 0.93) | 0.03 | 0.73 (0.29, 1.84) | 0.50 |
| Morbid obesity (BMI$\geq 40$ kg/m<sup>2</sup>) | | | | |
| No | Reference |  |  |  |
| Yes | 0.86 (0.29, 2.61) | 0.80 |  |  |
| Unknown | 1.11 (0.31, 3.97) | 0.88 |  |  |
| Heart diseases | 4.24 (2.03, 8.88) | <0.001 | 2.97 (1.01, 8.70) | **0.048** |
| Lung diseases | 1.04 (0.47, 2.32) | 0.92 |  |  |
| Hypertension | 3.52 (1.69, 7.34) | 0.001 | 0.77 (0.28, 2.09) | 0.61 |
| Diabetes | 1.16 (0.59, 2.29) | 0.66 |  |  |
| Immunocompromised conditions | 1.66 (0.59, 4.70) | 0.34 |  |  |
| Lymphopenia <1000 cells/mm<sup>3</sup> | 7.42 (3.02, 18.25) | <0.001 | 6.62 (2.19, 20.00) | **0.001** |
| Creatinine elevation >1.20 mg/dl | 5.98 (2.99, 12.01) | <0.001 | 2.36 (0.91, 6.08) | 0.08 |
| CRP>100 mg/dl |  |  |  |  |
| No | Reference |  | Reference |  |
| Yes | 2.83 (1.35, 5.93) | 0.006 | 1.78 (0.67, 4.68) | 0.25 |
| Unknown | 2.38 (0.46, 12.26) | 0.30 | 87.47 (1.52, 5038.05) | **0.03** |
| D-dimer>1000 ng/ml |  |  |  |  |
| No | Reference |  | Reference |  |
| Yes | 3.42 (1.56, 7.50) | 0.002 | 1.40 (0.49, 3.99) | 0.53 |
| Unknown | 1.97 (0.39, 10.03) | 0.42 | 0.15 (0.004, 6.51) | 0.33 |
| Troponin elevation within 72 hours | 3.68 (1.46, 9.27) | 0.006 | 0.91 (0.26, 3.23) | 0.89 |

### SARS-CoV-2 viremia at the time of ED presentation was associated with diffuse tissue damage, tissue fibrosis/repair and elevation of proinflammatory markers

We included in the proteomic analysis 247 participants with either viremia above quantification range (viremic) or undetectable viremia (aviremic). Unsupervised clustering of participants by UMAP using Olink proteomic results demonstrated a clear separation of the majority of viremic participants from aviremic participants (Figure 3A). In hierarchical clustering of participants by viremia-associated protein signatures, viremic participants were dispersed into several distinct clusters, indicating the heterogeneity of proteomic signatures among viremic participants (Supplementary Figure S6). In addition, viremia and severe disease showed overlap in the proteomic signatures (Supplementary Figure S6).

**Figure 3.**
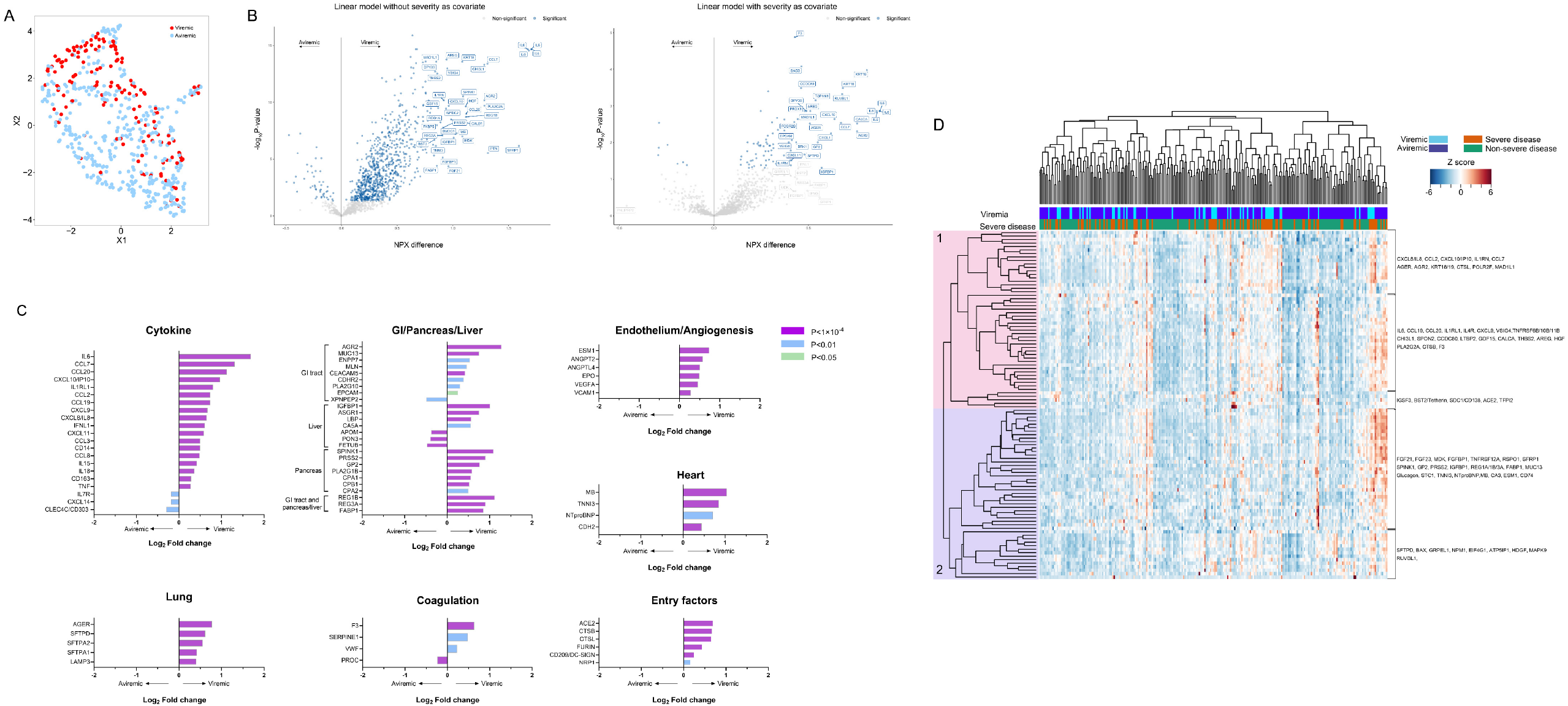
Plasma proteomic biomarkers and predictors of disease severity. (A) Unsupervised clustering UMAP for COVID-19-positive patients at days 0, 3 and 7. Red dots indicate viremic participants and blue dots indicate aviremic participants. (B) Volcano plots showing NPX differences in protein levels between viremic and aviremic participants. The left panel is derived from a linear model without severity as a covariate; the right panel is derived from a linear model with severity as a covariate. (C) Representative differentially expressed proteins in between viremic and aviremic participants. Adjusted P values are color coded as indicated. (D) Heatmap of the top 100 differentially expressed proteins between viremic and aviremic participants. Each row represents expression of an individual protein over the entire cohort; each cell represents the Z score of protein expression for all measurements across a row. Selected proteins are indicated.

To identify differentially expressed proteins between viremic and aviremic participants, we created linear models to fit each of the proteins at Day 0 with viremia status as a main effect and adjusted for age, demographics, and key comorbidities (Figure 3B). A number of prominent proteomic pathways were associated with higher plasma viral load. First, viremic participants demonstrated higher expression of viral response and interferon/monocytic pathway proteins including IL6, C-C Motif Chemokine Ligand 7 (CCL7)/monocyte-chemotactic protein 3 (MCP3), CCL20/macrophage inflammatory protein 3 alpha (MIP3A), CXCL10/Interferon gamma-induced protein 10 (IP-10), CXCL9/monokine induced by gamma interferon (MIG), CXCL8/IL8, interferon lambda 1 (IFNL1), CCL2/MCP1, CCL19/MIP3B, CCL3/MIP1A, CXCL11, IL15, and IL18 (Figure 3C). Nicotinamide phosphoribosyl transferase (NAMPT), an important regulator upstream to IL6 production^16^, was also upregulated in the viremic group. Second, viremia was associated with elevation of tissue damage markers^17^, including gastrointestinal (GI) tract/pancreas/liver markers (e.g. REG3A, REG1B, AGR2, GP2, MUC13, FABP1, PLA2G1B, PLA2G10, SPINK1, EPCAM, IGFBP1), lung markers especially surfactant proteins (SFTPD, SFTPA1/2, AGER, LAMP3), and cardiac markers (Troponin I3/TNNI3, NTproBNP, MB, CDH2). KRT18, KRT19, and RUVBL1 which are widely expressed in a variety of tissue types, including GI tract, pancreas, lungs, urinary system, and adipose tissue, are also significantly elevated in viremic participants, serving as markers of pan-tissue damage. It is worth mentioning that some of these proteins are also likely playing an important role in tissue fibrosis, including SERPINE1, CHI3L1, CTSL, along with TGF A/B and type IV collagen proteins (COL6A3, COL4A1). Third, higher plasma viral load was associated with signs of endovascular damage, with prominent endothelium/vascular markers and angiogenesis related proteins (ANGPT2, ANGPTL4, EPO, ESM1, VEGFA, VCAM), and coagulation pathway related markers (F3/tissue factor, SERPINE1, slight elevation of VWF, along with downregulation of PROC) (Figure 3C). In addition, we noted upregulation in viremic participants of certain complement pathway related proteins, especially PTX3, and to a lesser degree C1QA.

After adjusting for disease severity in the models, certain proinflammatory markers (IL6, CCL7, CXCL10/IP10, CXCL11), pulmonary injury markers (SFTPD, SFTPA1/2, AGER), GI tract/pancreas/liver markers (AGR2, IGFBP1, PLA2G10, EPCAM, MUC13, GP2), coagulation markers (F3), tissue fibrosis marker (CHI3L1) and pan-tissue injury markers (e.g. epithelial cell proteins RUVBL1, KRT18/19) remained significantly associated with SARS-CoV-2 viremia, independent of disease severity (Figure 3B). Interestingly, we also noted elevation of certain proteins that facilitate SARS-CoV-2 infection, including its receptor ACE2 ^18^, CD209/DC-SIGN ^19^, NRP1 ^20,21^, and entry facilitators/proteases FURIN ^22^, Cathepsin B/L (CTSB/CTSL)^23^ (Figure 3C). Lactate dehydrogenase (LDH), a commonly used laboratory marker indicating tissue damage and pyroptosis ^24^, was highly correlated to lung-related, severity independent markers (SFTPA 1/2, AGER), especially in the viremic group (Supplementary Figure S7).

In addition to proteins related to tissue injury, fibrosis and repair, we noted significant elevation of certain monocytes/dendritic cells (i.e., CD14, CD163) and plasmablasts (i.e. CD138/SDC1, TXNDC5) related proteins based on PBMC gene expression/RNA database ^25-28^. Certain neutrophil markers were also elevated in viremic group, including CHI3L1, IL1RN, MMP9, and PRTN3 (Proteinase-3)^28^ (Supplementary Table S3). After adjusting for severe disease, certain monocytes/dendritic cells markers and neutrophil markers remained significantly associated with viremia (Supplementary Table S3).

To further dissect the relationship of viremia-associated differentially expressed proteins, we again performed unsupervised hierarchical clustering of participants by viremia-associated protein from Day The top 100 differentially expressed proteins from the linear model were clustered in Figure 3D. In Cluster 1, IFN-I and monocyte-related cytokines and proteins were grouped together (including IL6, CXCL10 etc.) in addition to neutrophil-related (CHI3L1), and NK cell related (SPON2) proteins ^26,28^. SPON2 ^29^ and PLA2G2A ^30^ from this cluster also play a role in innate immune response. Certain tissue related proteins including lung (AGER), GI (AGR2), epithelial cells markers (KRT18 and KRT19), tissue factor (F3), tissue repair/growth related proteins (HGF, GDF15, CCDC80) and entry-related factors (ACE2, CTSL, and CTSB) were also found in this cluster. In comparison, Cluster 2 included several tissue repair/fibroblast-related proteins, heart and skeletal muscle, GI tract and pancreas-related proteins. Finally, SFTPD, a locally secreted surfactant protein in lungs, clustered with certain apoptosis-related proteins (i.e., BAX) and housekeeping proteins located in the cytosol (NPM1, MAPK9, EIF4G1) and mitochondria (ATP5IF1, GRPEL1). Lung tissue markers, including SFTPA1/2 and AGER, were moderately correlated to upstream apoptosis-related protein (Fas, PDCD family and BAX/BID/BCL2L11) and weakly correlated to pyroptosis-related proteins (Supplementary Figure S8). Using elastic-net logistic regression with cross-validation, SARS-CoV-2 viremia along with Day 0 proteomic data yielded good predictive performance for severe disease (AUC 0.83, 95% CI 0.80, 0.86; Supplementary Figure S5).

### Viremic participants experienced prolonged tissue damage, inflammation, and elevation in viral entry factors

To assess the longitudinal impact of viremia, we focused on 103 hospitalized participants with complete proteomic data from Day 0, 3 and 7 (acuity level from A1 to A4). We first looked at the trajectory of those proteins identified in the Day 0 analysis (Figure 3). Viremic participants had persistently higher levels of proinflammatory markers beyond day 0, especially those related to monocyte activation. For some inflammatory markers (e.g., TNF, IL18, and CD14), differences between groups became highly divergent over time with hyper-accentuated inflammatory responses in viremic participants (Figure 4A). Longitudinal proteomic analysis also demonstrated the persistent elevation of proteomic pathways reflecting organ damage, endothelial damage, and a hypercoagulable state. Certain complement pathway related proteins and entry-related factors were also persistently elevated in the viremic group (Figure 4A).

**Figure 4.**
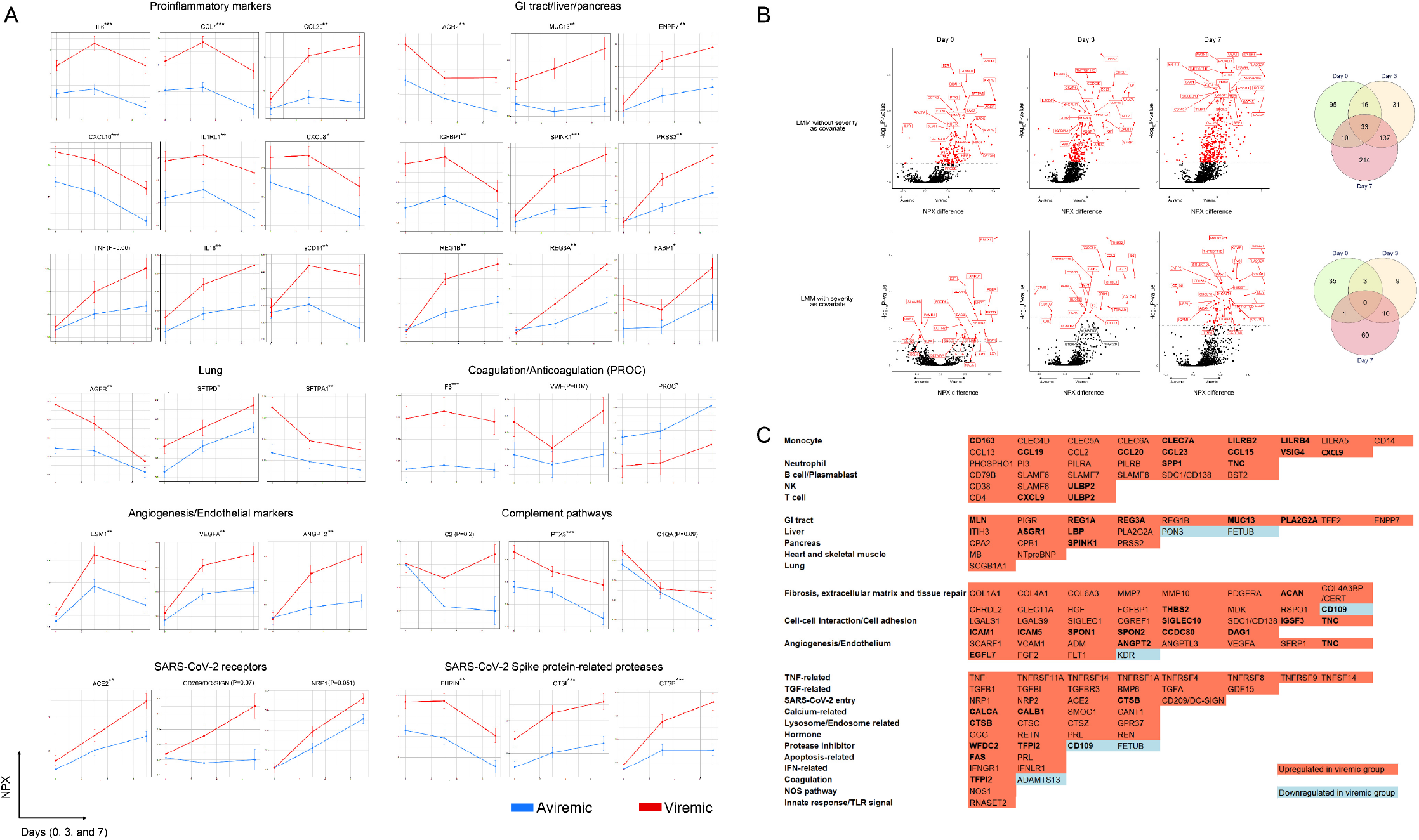
Temporal trends of differentially expressed proteins between viremic and aviremic groups. (A) Point range plots of differentially expressed proteins between viremic and aviremic groups, only including patients with samples on all days 0, 3, and 7. Error bar is standard error of mean (SEM). P value of viremia was derived from a linear mixed model (LMM) accounting for the interaction between time points and viremia; *, P<0.05; **, P<0.01; ***, P<0.001. (B) Volcano plots showing LMM of differentially expressed proteins at different time points (P values indicate group differences calculated by the Tukey posthoc method). Venn diagrams demonstrate the overlap of differentially expressed proteins at different time points. (C) Selected proteins differentially expressed in the viremic group later in hospitalization (only at Day 7 or only at Day 3+Day 7). Bold font indicates statistical significance after adjusting for severe disease.

We next fit linear mixed models (LMMs) for each protein with time and viremia status as main effects and adjusted for age, demographics, and key comorbidities to identify proteins that were significant for the interaction between viremia and time (Figure 4B). We further noted an uptrend in monocyte-related proteins in the viremic group at later time points, followed by neutrophil and B-cell/plasmablast related proteins (Figure 4C). Many of these markers were significantly elevated even after adjustment for severe disease (labeled in bold font). We also noted an association between viremia and persistent, yet uptrending tissue damage levels, especially those from GI system. Furthermore, levels of numerous tissue fibrosis/tissue repair/extracellular matrix related proteins began to increase at later time points, including several collagen proteins, ACAN, MDK etc. (Figure 4C). In parallel, endothelial damage and angiogenesis-related proteins were further upregulated in the viremic group, in conjunction with a dysregulated hemostasis state featured by decreases in ADAMTS13 (Figure 4A and 4C).

### Viremia at the time of ED presentation is not associated with neutralizing antibody levels

Finally, we evaluated the relationship between SARS-CoV-2 viremia and neutralization level. We included participants with neutralization data available at baseline and at least one follow-up time point. Neutralization levels between viremic and aviremic groups were not significantly different at days 0, 3, and 7 (Figure 5A). In the subset of participants with neutralization data available beyond day 7, no clear difference was observed between viremic and aviremic groups (Figure 5B). At the time of ED presentation, levels of SDC1/CD138, a cardinal and specific marker for plasmablasts^26,28^, was significantly correlated with neutralization level, irrespective of the presence of viremia (Figure 5C). We also conducted an analysis including a subgroup of participants with available viral load at Day 3 (n=49) and Day 7 (n=39). Undetectable viral load at Day 3 or Day 7 was not associated with higher neutralizing antibody titers (Supplementary Figure S9).

**Figure 5.**
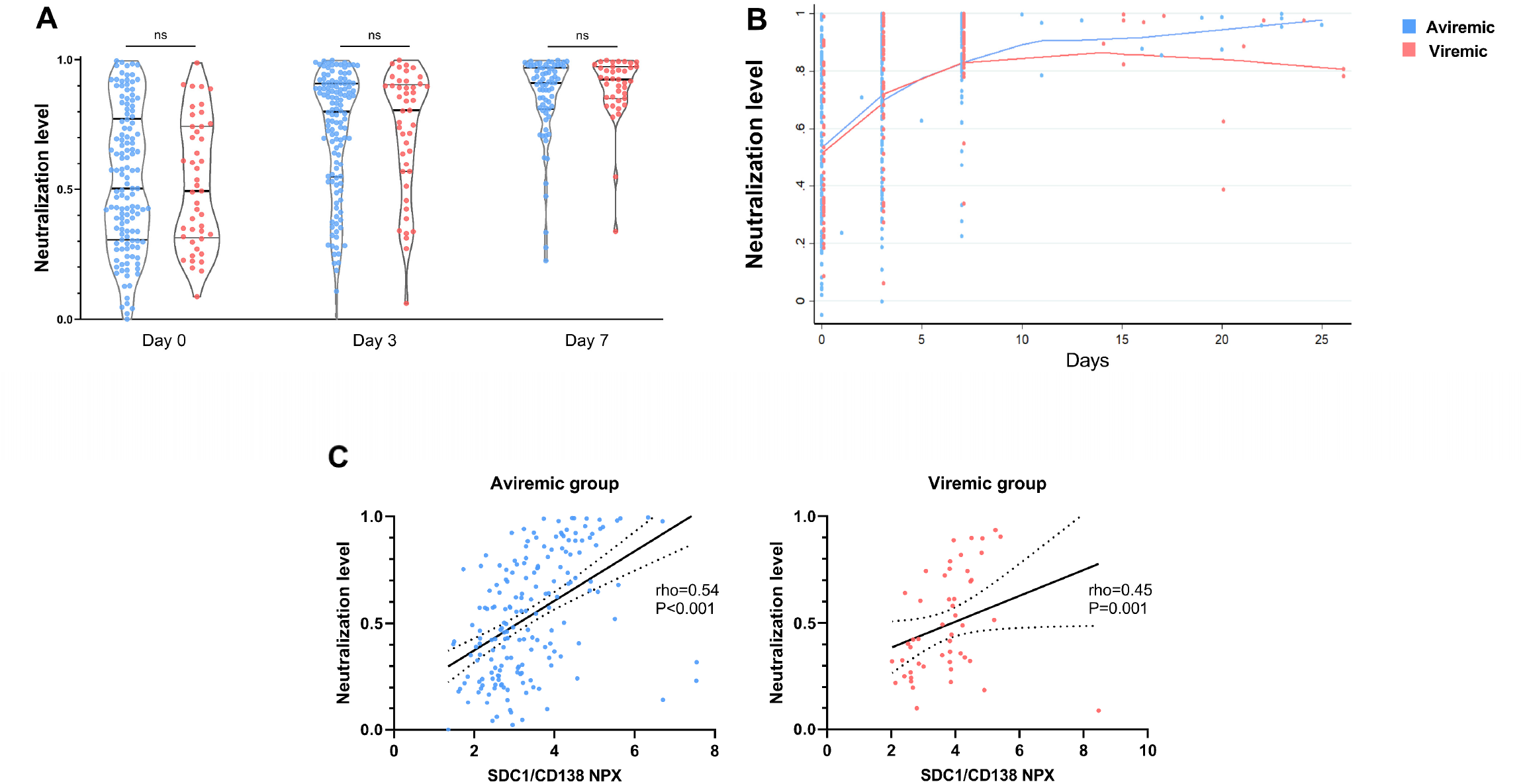
Neutralization level and viremia. (A) Violin plot of neutralization levels stratified by viremia status. Mann–Whitney U test was used to evaluate the difference between two groups. ns, not significant. (B) Neutralization rate between viremic and aviremic groups. Lowess smooth regression was performed to depict the trajectory of neutralizing rates between two groups. (C) Correlation between SDC1/CD138 (a marker for plasmoblast) NPX and neutralizing rate at Day 0. Linear regression (solid line) with 95% confidence intervals (dotted lines) are shown. Spearman correlation was used to evaluate the correlation between SDC1/CD138 NPX and neutralizing rates.

## Discussion

In this study, we report a comprehensive analysis of SARS-CoV-2 viremia and its associations with disease outcomes and proteomic pathways from a cohort of ED patients with COVID-19. To our knowledge, this is the largest longitudinal cohort to explore this topic. The results demonstrate that SARS-CoV-2 plasma viremia at the time of ED presentation predicts maximal COVID-19 disease severity and mortality within 28 days. In addition, we for the first time uncovered proteomic signatures upregulated in the setting of SARS-CoV-2 viremia, including prominent pathways highlighting lung and systemic tissue damage, tissue fibrosis and repair, a pronounced proinflammatory response, perturbed hemostasis, and upregulation of viral entry factors.

It is now clear that SARS-CoV-2 infection extends outside the respiratory system^2^, and the detection of plasma viremia represents the “link” for extrapulmonary multiorgan involvement and adverse outcomes. Systemic invasion from the respiratory tract is not unique to SARS-CoV-2, as viremia has also been described for other respiratory viruses including SARS-CoV-1^31^, influenza virus^32^, respiratory syncytial virus^33^, and adenovirus^34^. We and others have previously demonstrated that SARS-CoV-2 viremia is more commonly detected in critically ill populations^11,12,14,35^, and is correlated with cardinal proinflammatory markers, including IL6 ^11,14^, IP10/CXCL10 ^36^, CCL2/MCP1 ^36^ and markers of endothelial damage ^36^. These studies were limited by a lack of true viral load quantification, small sample sizes that could not account for confounders, and/or the evaluation of hospitalized patients only late in their disease course. Here, we report the largest study to date of plasma SARS-CoV-2 plasma viremia using a quantitative viral load assay that allowed for the confirmation of the previous findings ^11,14^ even after adjustment of multiple potential confounding variables. A particular strength of our study was the ability to enroll all acutely ill patients upon ED arrival and thereby minimize selection bias. Our results demonstrate that at the time of ED presentation, plasma SARS-CoV-2 viral load levels independently predicted, in a dose-dependent manner, severe disease and death within the next 28 days. SARS-CoV-2 viremia was associated with clinical markers associated with disease severity, including elevated CRP and lymphopenia.

Our proteomic analysis represents another strength of this study, which demonstrates unique pathways in patients with plasma viremia that together orchestrate a “perfect storm”. Viremic individuals displayed a proteomic pattern of broad tissue damage, highlighted by severe lung damage, GI damages, persistent proinflammatory markers elevation, endovascular damage, and tissue fibrosis. While previous studies have reported the elevation of certain nonspecific tissue damage markers in viremic individuals, especially LDH^36,37^, our study allows a far more precise evaluation and demonstrates that respiratory tract and GI tract/liver/pancreas injuries constitute some of the major contributors to tissue injury in patients with SARS-CoV-2 viremia. Our proteomic analysis extend the results of a proteomic evaluation of an autopsy tissue study^38^ by showing that many of the pathways of tissue and endothelial cell damage can already be identified relatively early in the disease course and may be mediated by systemic dissemination of SARS-CoV-2 infection.

We observed the upregulation of a panel of angiogenesis and endothelial damage-related markers in viremic patients. In addition, several key factors in the coagulation pathway, including Factor III (F3), von Willebrand factor (VWF), SERPINE1 (plasmin inhibitor), were elevated in the viremic group, in conjunction with a decrease in ADAMTS13, a metalloprotease enzyme that cleaves and inhibits the activity of VWF. The presence of endothelial cell damage and dysregulation of the coagulation cascade is consistent with results in patients with critical disease or after death^38-40^. Our results not only demonstrate that these pathways become altered even in patients with early disease, but also provides the mechanistic link between plasma viremia and the hypercoagulable state observed in patients across the spectrum of COVID-19 disease severity^2,41,42^. These findings suggest that early interventions to prevent the circulatory dissemination of SARS-CoV-2 infection could help prevent these potentially devastating complications of COVID-19.

Interestingly, our proteomic analysis also showed a relationship between higher levels of viremia and persistent elevation of several SARS-CoV-2 entry factors, including FURIN and Cathepsin B/L (CTSB/CTSL). These results are consistent with reports of a proteomic analysis of COVID-19 autopsies ^38^ in which Nie and colleagues reported that CTSB/CTSL, which are proteases facilitating viral entry, are prominently elevated in the lungs. In contrast, ACE2, the primary host receptor for SARS-CoV-2, showed minimal upregulation^38^. Cathepsin family proteins are also known for their role in facilitating SARS-CoV-2 spike protein priming as well as promoting the inflammasome/pyroptosis pathway, as the autopsy studies also reveal that inflammasome/pyroptosis-related proteins, including LDH and MPO, are highly upregulated in lung tissues ^38^. The combination of our results and these autopsy findings point to the crucial role of the Cathepsin family proteins in the pathogenesis of SARS-CoV-2.

Of note, the viremic and aviremic groups had comparable neutralization activity in this cohort. Most patients presented fairly early during their symptom course and it is possible that neutralizing antibody titers were not yet particularly high or effective. More robust neutralizing antibody levels were detected during the course of hospitalization for all participants, regardless of initial level of plasma viremia. It could also be that the level of plasma viremia is a reflection of the extent of tissue-based infection and less a reflection of the current level of neutralizing antibody titers.

Our study also has a few notable limitations. Although quite comprehensive, our proteomic database does not cover all the cytokines and proteins of interest in COVID-19 pathogenesis. We rely on a pre-existing proteomic database ^17^ and peripheral blood databases ^26,28^ to infer the origin of differentially expressed proteins, but do not have data on scRNA-Seq from this cohort to confirm the cellular source of some differentially expressed protein. Given the relatively high limits of detection of culture-based assays, we are unable to confirm whether the RNA detected in plasma samples are from viable, infective SARS-CoV-2 virions.

In summary, we report the largest study to date that demonstrates SARS-CoV-2 viremia predicts severe COVID-19 disease outcomes and the likely role of systemic viral dissemination in mediating tissue damage, tissue fibrosis, hypercoagulable state, persistent elevation of proinflammatory markers, and higher viral entry factor expression. Our findings provide key insights into SARS-CoV-2 pathogenesis and identify potential therapeutic targets to mitigate COVID-19 disease severity.

## Methods

### Study participants

Participant enrollment was described in our prior report ^15^. Briefly, participants were enrolled in the Emergency Department (ED) from Massachusetts General Hospital, Boston MA, from 3/24/2020 to 4/30/2020 during the first peak of the COVID-19 surge, with an institutional IRB-approved waiver of informed consent. Symptomatic participants of 18 years or older with nucleic acid tests confirmed of SARS-CoV-2 infection were included in this current study. Clinical course was followed to 28 days post-enrollment, or until hospital discharge if that occurred after 28 days.

Enrolled participants who were SARS-CoV-2 positive (N=306) were categorized into five outcome/acuity groups: 1) A1, Death within 28 days, 2) A2, Requiring mechanical ventilation and survival to 28 days, 3) A3, Requiring hospitalization on supplemental oxygen within 28 days, 4) A4, Requiring hospitalization without needing supplemental oxygen, and 5) A5, Discharge from ED and not subsequently requiring hospitalization within 28 days. Severe disease was defined as belonging to group A1 or A2. In this current analysis, we only included participants with available plasma SARS-CoV-2 viral load (n=300).

### Study endpoints

The primary endpoint of this study is severe COVID-19 within 28 days of enrollment (intubation and/or death). Secondary endpoints include 28-day mortality and SARS-CoV-2 viremia.

### Plasma SARS-CoV-2 viral load

Plasma SARS-CoV-2 viral load measurement was reported in our previous study ^11^ with the following modifications. Briefly, RNA was extracted from 300μL of RPMI-1640 diluted ethylenediaminetetraacetic acid (EDTA)-preserved plasma sample (RPMI-1640: Plasma 2:1 dilution)^15^ using TRIzol™-based method (Thermo Fisher Scientific, Waltham, MA). SARS-CoV-2 viral load was quantified using the US CDC 2019-nCoV_N1 primers and probe set ^11^. The lower limit of SARS-CoV-2 N gene quantification was 100 copies/mL. Samples with a positive signal but viral load <100 copies/mL were denoted as detectable but unquantifiable.

### Olink proteomic analyses

Proteomic analyses were described in a prior report ^15^. Briefly, The Olink Proximity Extension Assay (PEA) is a technology developed for high-multiplex analysis of proteins. Oligonucleotide-labelled monoclonal or polyclonal antibodies (PEA probes) are used to bind target proteins in a pair-wise manner thereby preventing all cross-reactive events. Upon binding, the oligonucleotides come in close proximity and hybridize followed by extension generating a unique barcode for identification. The full Olink library contains 1472 proteins and 48 controls assays, dividing into inflammation, oncology, cardiometabolic and neurology panels, with overlap in interleukin (IL)6, IL8/C-X-C motif chemokine ligand (CXCL8), and tumor necrosis factor (TNF)-alpha for quality control (QC) purpose. Level of proteins were denoted as normalized protein expression (NPX) units through a QC and normalization process developed and provided by Olink. Data generation of NPX consists of normalization to the extension control (known standard), log2-transformation, and level adjustment using the plate control (plasma sample). Information regarding protein expression at tissue and blood cells levels, protein function, and protein localization was derived from the Human Protein Atlas ^43,44^.

### SARS-CoV-2 S pseudotyped lentivirus generation

Neutralizing antibody level was evaluated by pseudotyped lentivirus neutralization assay as reported in our recent study ^15^. Lentivirus vector was constructed using PCR amplification (Q5 High-Fidelity 2X Master Mix, New England Biolabs) from pUC57-nCoV-S (gift of Jonathan Abraham), in which the C-terminal 27 amino acids of SARS-CoV-2 S are replaced by the NRVRQGYS sequence of HIV-1 ^45^. The truncated SARS-CoV-2 S fused to gp41 was cloned into pCMV by Gibson assembly to obtain pCMV-SARS2ΔC-gp41. Other vectors including psPAX2, pCMV-VSV-G, pTRIP-SFFV-EGFPNLS (Addgene plasmid #86677), and pTRIP-SFFV-Hygro-2ATMPRSS2 were described in our recent publication ^15^. 293T ACE2/TMPRESS2 cell line was generated as described in our recent publication ^15^. 293T cells were seeded at 0.8 × 10^6^ cells per well in a 6-well plate and were transfected the same day with a mix of DNA containing 1 μg psPAX, 1.6 μg pTRIP-SFFV-EGFP-NLS, and 0.4 μg pCMV-SARS2ΔC-gp41 using TransIT^®^-293 Transfection Reagent. After overnight incubation, the medium was changed. SARS-CoV-2 S pseudotyped lentiviral particles were collected 30-34 hours post medium exchange and filtered using a 0.45 μm syringe filter. To transduce 293T ACE2 cells, the same protocol was followed, with a mix containing 1 μg psPAX, 1.6 μg pTRIPSFFV-Hygro-2A-TMPRSS2, and 0.4 μg pCMV-VSV-G.

### SARS-CoV-2 S pseudotyped lentivirus antibody neutralization assay

One day before neutralization experiment, 293T ACE2/TMPRSS2 cells were seeded at 5 × 10^3^ cells in 100 μl per well in 96-well plates. On the day of lentiviral harvest, 100 μl SARS-CoV-2 S pseudotyped lentivirus was incubated with 50 μl of plasma diluted in medium to a final concentration of 1:100. Medium was then removed from 293T ACE2/TMPRSS2 cells and replaced with 150 μl of the mix of plasma and pseudotyped lentivirus. Wells in the outermost rows of the 96-well plate were excluded from the assay. After overnight incubation, medium was changed to 100 μl of fresh medium. Cells were harvested 40-44 hrs post infection with TrypLE (Thermo Fisher), washed in medium, and fixed in FACS buffer containing 1% PFA (Electron Microscopy Sciences). Percentage GFP was quantified on a Cytoflex LX (Beckman Coulter), and data was analyzed with FlowJo. Neutralization rate was defined as 1- (GFP%_pseudovirus+plasma_/GFP%_pseudovirus alone_).

## Statistics

We summarized continuous variables using median and interquartile ranges (IQRs). For clinical variables, we used the Wilcoxon rank-sum test to compare continuous variables from two different categorical groups and Dunn’s test for three or more groups. Categorical variables were evaluated using the χ^2^ test or Fisher’s exact test. We used Spearman’s rank correlation coefficient to evaluate correlation between different continuous variables. To evaluate the association of plasma SARS-CoV-2 viral load and clinical outcomes, we used logistic regression analyses to calculate odds ratio (OR) and 95% confidence intervals (CI). Both univariate and multivariate logistic regression analyses were performed. In multivariate analyses, factors with a P value <0.10 from univariate models were included. We also used Cox proportional model to evaluate the correlation between viremia and 28-day mortality by calculating the hazard ratio (HR). Clinical data analyses, logistic regression and Cox proportion regression were performed on Stata (version 13.1) and figures were generated by Stata and GraphPad (Prism, version 9.0). R (version 4.0.2) was used to analyze proteomic data.

### Linear models

Linear regression models were fit independently to each protein using the lm package in R with protein values (NPX for Olink data) as the dependent variable. The models included a term for viremia and covariates for age, sex, ethnicity, heart disease, diabetes, hypertension, hyperlipidemia, pulmonary disease, kidney disease, immunocompromised status to control for any potential confounding. P-values were adjusted to control the false discovery rate (FDR) at 5% using the Benjamini-Hochberg method implemented in the emmeans package in R.

### Linear mixed models

Linear mixed effects models (LMMs) were fit independently to each protein using the lme4 package in R with protein values (NPX for Olink data) as the dependent variable. The model for viremia included a main effect of time, a main effect of viremia, the interaction between these two terms, and a random effect of patient ID to account for the correlation between samples coming from the same patient. Covariates for age, sex, ethnicity, heart disease, diabetes, hypertension, hyperlipidemia, pulmonary disease, kidney disease, and immuno-compromised status were included in the model to control for any potential confounding effects. Details were reported in our recent study^15^.

## Data Availability

Data will be available once the manuscript is formally accepted after peer-review.

## Acknowledgments

We want to thank all the participants in this study. We thank the all the clinical staff who made sample collection possible.

Direct funding for this project was provided in part by a grant from Mark, Lisa and Enid Schwartz (to J.Z.L.), the National Institute of Health (N.H., U19AI082630), an American Lung Association COVID-19 Action Initiative grant (M.B.G.), and grants from the Executive Committee on Research at MGH (M.B.G. and M.R.F.), the Chan-Zuckerberg Initiative (A-C.V.). N.H. was also funded by a gift from Arthur, Sandra and Sarah Irving for the David P. Ryan, MD Endowed Chair in Cancer Research. M.G. is the recipient of an EMBO Long-Term Fellowship (ALTF 486-2018) and a Cancer Research Institute/Bristol-Myers Squibb Fellow (CRI2993). This work was also supported by the Harvard Catalyst / Harvard Clinical and Translational Science Center (National Center for Advancing Translational Sciences, National Institutes of Health Awards UL1 TR 001102 and UL1 TR 002541-01) and by the Harvard University Center for AIDS Research (NIAID 5P30AI060354). We are also very grateful for the generous contributions of Olink Proteomics Inc. and Novartis (in collaboration with SomaLogic,Inc.) for providing in-kind all proteomics assays presented in this work, without which our findings would not have been possible.

## Author Contributions

Conceptualization: JZL, YL

Establishment of the MGH ED cohort: MRF, BAP, AV, MS-F, NH, MBG

Resources: MRF, NH, MBG

SARS-CoV-2 viral load assay: YL, JR, JPF

Neutralization assay: MG

Sample collection: NC, AG, IG, HK, TL, KL, BL, CL, KM, JM, BM, BAP, MR-L, BR, NS, JT, MT

Formal Analysis: YL, AS, AM

Writing – Original Draft: YL

Writing – Review & Editing: JZL, BAP, MRF, AS, AM, NH, MBG, JR, JPF

