## Supplementary Materials for "SARS-CoV-2 Viremia is Associated with Distinct Proteomic Pathways and Predicts COVID-19 Outcomes"

### Supplementary Methods

##### Unsupervised clustering

Principal components analysis (PCA) was performed using all proteins and all samples using the prcomp() function in R. Unsupervised clustering by UMAP was performed using all proteins, and either all samples or just Day 0 samples, using the umap() function in R. UMAP coordinates were plotted using the ggplot2 package.

### Supplementary Tables

#### Supplementary Table S1. Association of SARS-CoV-2 viral load and clinical outcomes, sensitivity analyses.

| | Univariate OR (95% CI) | P | Multivariate OR (95% CI) | P |
|---|---|---|---|---|
| **Severe diseases[a]** | | | | |
| **SARS-CoV-2 viral load accounting for detectable but non-quantifiable viremia** | | | | |
| Aviremic | Reference | | Reference | |
| Viremic but below quantification range | 1.91 (0.99, 3.66) | 0.054 | 2.01 (0.91, 4.42) | 0.08 |
| Viremic ≥ 2 log copies/ml | 14.66 (6.82, 31.53) | <0.001 | 11.73 (4.79, 28.74) | <0.001 |
| **SARS-CoV-2 viral load as continuous variable** | | | | |
| Viral load (per 1 log copy/ml increase) | 2.57 (1.93, 3.43) | <0.001 | 2.49 (1.75, 3.54) | <0.001 |
| **Death at day 28[b]** | | | | |
| **SARS-CoV-2 viral load accounting for detectable but non-quantifiable viremia** | | | | |
| Aviremic | Reference | | Reference | |
| Viremic but below quantification range | 1.58 (0.62, 4.05) | 0.34 | 1.17 (0.36, 3.83) | 0.79 |
| Viremic ≥ 2 log copies/ml | 4.92 (2.30, 10.53) | <0.001 | 4.01 (1.46, 11.06) | 0.007 |
| **SARS-CoV-2 viral load as continuous variable** | | | | |
| Viral load (per 1 log copy/ml increase) | 1.67 (1.33, 2.10) | <0.001 | 1.46 (1.08, 1.97) | 0.01 |

a, multivariate models were adjusted for age groups, heart diseases, lung diseases, hypertension, diabetes, immunocompromised conditions, lymphopenia, creatinine elevation, C reactive protein elevation, D-dimer elevation and troponin elevation. Seen in Table 2.

b, multivariate models were adjusted for age groups, race, heart diseases, hypertension, lymphopenia, creatinine elevation, C reactive protein elevation, D dimer elevation, troponin elevation. Seen in Table 2.

#### Supplementary Table S2. Clinical factors associated with viremia.

| | **Univariate OR (95% CI)** | **P** | **Multivariate OR (95% CI)** | **P** |
|---|---|---|---|---|
| Age | | | | |
| <50 years | Reference | | Reference | |
| 50-64 years | 1.82 (0.77, 4.29) | 0.17 | 1.33 (0.53, 3.34) | 0.54 |
| ≥65 years | 2.68 (1.23, 5.84) | 0.01 | 1.75 (0.69, 4.44) | 0.24 |
| Female (male as reference) | 0.60 (0.33, 1.11) | 0.10 | | |
| People of color (Caucasian as reference) | 1.52 (0.83, 2.76) | 0.18 | | |
| Morbid obesity (BMI≥40 $kg/m^2$) | | | | |
| No | Reference | | | |
| Yes | 1.24 (0.51, 3.04) | 0.64 | | |
| Unknown | 0.51 (0.11, 2.29) | 0.38 | | |
| Heart diseases | 0.66 (0.26, 1.65) | 0.37 | | |
| Lung diseases | 0.42 (0.17, 1.02) | 0.06 | 0.40 (0.15, 1.07) | 0.07 |
| Hypertension | 1.29 (0.71, 2.33) | 0.41 | | |
| Diabetes | 2.34 (1.28, 4.27) | 0.006 | 2.15 (1.13, 4.10) | **0.02** |
| Immunocompromised conditions | 1.18 (0.42, 3.31) | 0.75 | | |
| Lymphopenia <1000 $cells/mm^3$ | 1.86 (1.01, 3.43) | 0.045 | 1.38 (0.70, 2.71) | 0.35 |
| Creatinine elevation >1.20 mg/dl | 2.01 (1.04, 3.88) | 0.04 | 1.29 (0.58, 2.87) | 0.53 |
| CRP>100 mg/dl | | | | |
| No | Reference | | Reference | |
| Yes | 3.22 (1.66, 6.25) | 0.001 | 2.29 (1.10, 4.75) | **0.03** |
| Unknown | 0.83 (0.10, 6.93) | 0.86 | 0.41 (0.02, 8.45) | 0.47 |
| D-dimer>1000 ng/ml | | | | |
| No | Reference | | Reference | |
| Yes | 1.98 (1.05, 3.75) | 0.04 | 1.12 (0.54, 2.32) | 0.76 |
| Unknown | 0.97 (0.20, 4.67) | 0.98 | 2.36 (0.24, 23.28) | 0.46 |
| Troponin elevation within 72 hours | 2.57 (1.04, 6.36) | 0.04 | 1.90 (0.64, 5.68) | 0.25 |

Supplementary Table S3. Differentially expressed proteins related to peripheral blood cells.

List of protein derived from Monaco et al., Cell Rep 2019 [1].

| Protein | NPX difference (Viremic- Aviremic) | Adjusted P value (without severity as covariate) |
|---|---|---|
| | **Monocytes** | |
| **IL6** | 1.71 | 2.12E-15 |
| **CCL7** | 1.31 | 4.00E-14 |
| CCL20 | 1.12 | 1.06E-09 |
| **CXCL10** | 0.96 | 1.99E-10 |
| **VSIG4** | 0.95 | 1.06E-13 |
| **CTSL** | 0.65 | 2.80E-13 |
| PTX3 | 0.61 | 3.61E-09 |
| CD300E | 0.59 | 4.00E-08 |
| **CXCL11** | 0.58 | 3.59E-06 |
| PVR | 0.55 | 1.81E-10 |
| VCAN | 0.52 | 2.10E-07 |
| CLEC5A | 0.52 | 7.25E-09 |
| CCL3 | 0.50 | 4.30E-06 |
| CD14 | 0.50 | 8.22E-07 |
| CLEC4D | 0.50 | 3.81E-05 |
| CCL8 | 0.48 | 4.89E-05 |
| VMO1 | 0.45 | 9.60E-05 |
| SIGLEC10 | 0.44 | 5.34E-08 |
| CXCL16 | 0.43 | 4.09E-08 |
| CES1 | 0.42 | 1.18E-03 |
| HMOX1 | 0.40 | 2.13E-04 |
| **LRP1** | 0.40 | 1.31E-09 |
| **SIGLEC1** | 0.39 | 1.00E-07 |
| NID1 | 0.39 | 1.09E-07 |
| CLEC1B | 0.38 | 1.10E-02 |
| TNFRSF8 | 0.37 | 4.84E-05 |
| MSR1 | 0.36 | 4.37E-06 |
| CD300LF | 0.36 | 1.41E-02 |
| CCL13 | 0.36 | 4.50E-04 |
| TYMP | 0.35 | 6.65E-04 |
| SPINT1 | 0.31 | 3.22E-06 |
| CST3 | 0.30 | 2.30E-03 |
| CD163 | 0.29 | 2.50E-05 |
| CD93 | 0.28 | 1.52E-05 |
| LILRA5 | 0.27 | 5.57E-06 |
| PLXNB2 | 0.26 | 1.88E-06 |
| **OSCAR** | 0.24 | 1.91E-04 |
| CD302 | 0.22 | 3.73E-02 |
| SLAMF8 | 0.21 | 1.77E-03 |
| HBEGF | 0.21 | 2.34E-02 |
| LILRB2 | 0.20 | 3.71E-04 |
| C1QA | 0.18 | 1.37E-02 |
| CPVL | 0.18 | 3.48E-02 |
| | **Neutrophils** | |
| **CHI3L1** | 1.23 | 4.82E-14 |
| **IL1RN** | 0.92 | 6.62E-11 |
| ADM | 0.73 | 1.91E-10 |
| CD177 | 0.51 | 1.18E-05 |
| **TNFSF13B** | 0.50 | 1.54E-08 |
| S100A12 | 0.48 | 3.66E-09 |
| VNN2 | 0.45 | 1.20E-05 |
| SIRPB1 | 0.41 | 5.11E-06 |
| PRTN3 | 0.41 | 9.02E-07 |

| | | |
|---|---|---|
| S100P | 0.37 | 5.76E-04 |
| SIGLEC5 | 0.37 | 1.50E-02 |
| LCN2 | 0.35 | 6.28E-04 |
| MMP9 | 0.28 | 4.88E-03 |
| DEFA1 | 0.24 | 3.28E-02 |
| **Plasmablasts** | | |
| CD138/SDC1 | 0.83 | 4.73E-06 |
| RRM2 | 0.59 | 6.86E-07 |
| MZB1 | 0.46 | 1.09E-04 |
| TXNDC5 | 0.44 | 1.71E-04 |
| **CD4+T cells** | | |
| TNFRSF4 | 0.26 | 7.89E-03 |
| **NK cells** | | |
| NCR1 | 0.17 | 2.35E-02 |

Bold font indicates statistical significance after further adjustment for severity. Of note, NPX is already a $Log_2$ transformed index.

#### Supplementary Figures

Supplementary Figure S1. Correlation between viral load and duration between symptom onset and ED presentation.

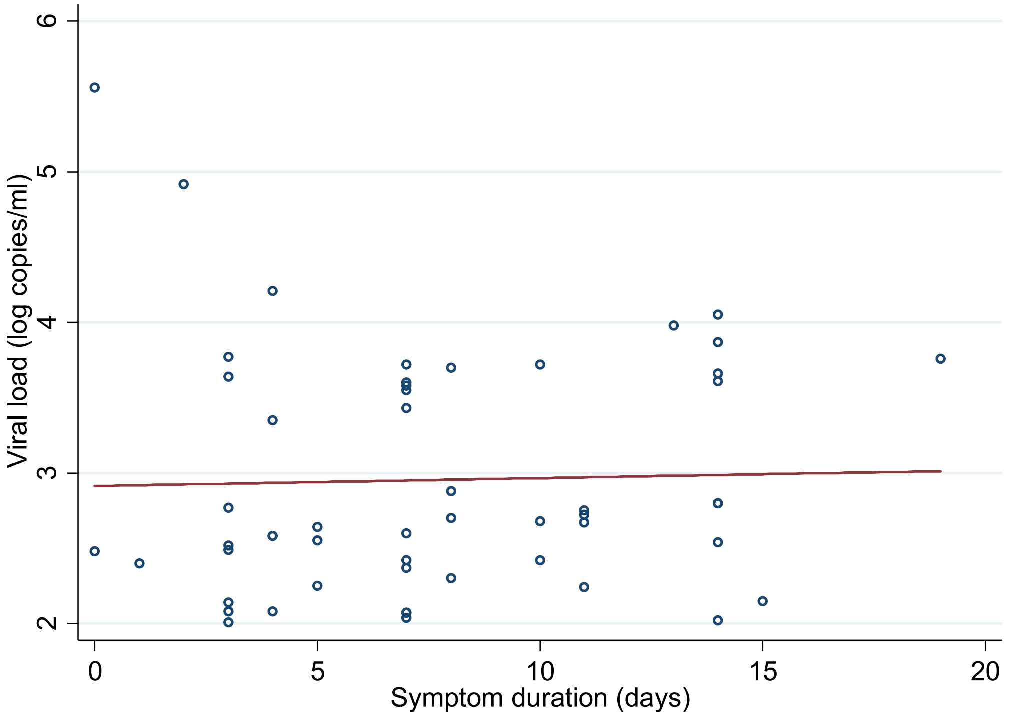

Supplementary Figure S2. Snapshots of disease severity at Day 0, 3, 7 and 28.

The left column was grouped by quantifiable viremia (≥ 2 log copies/ml) and undetectable or unquantifiable viremia (< 2 log copes/ml). The right column was grouped by quantifiable viremia, unquantifiable viremia (detectable but < 2 log copies/ml) and undetectable viremia (aviremic).

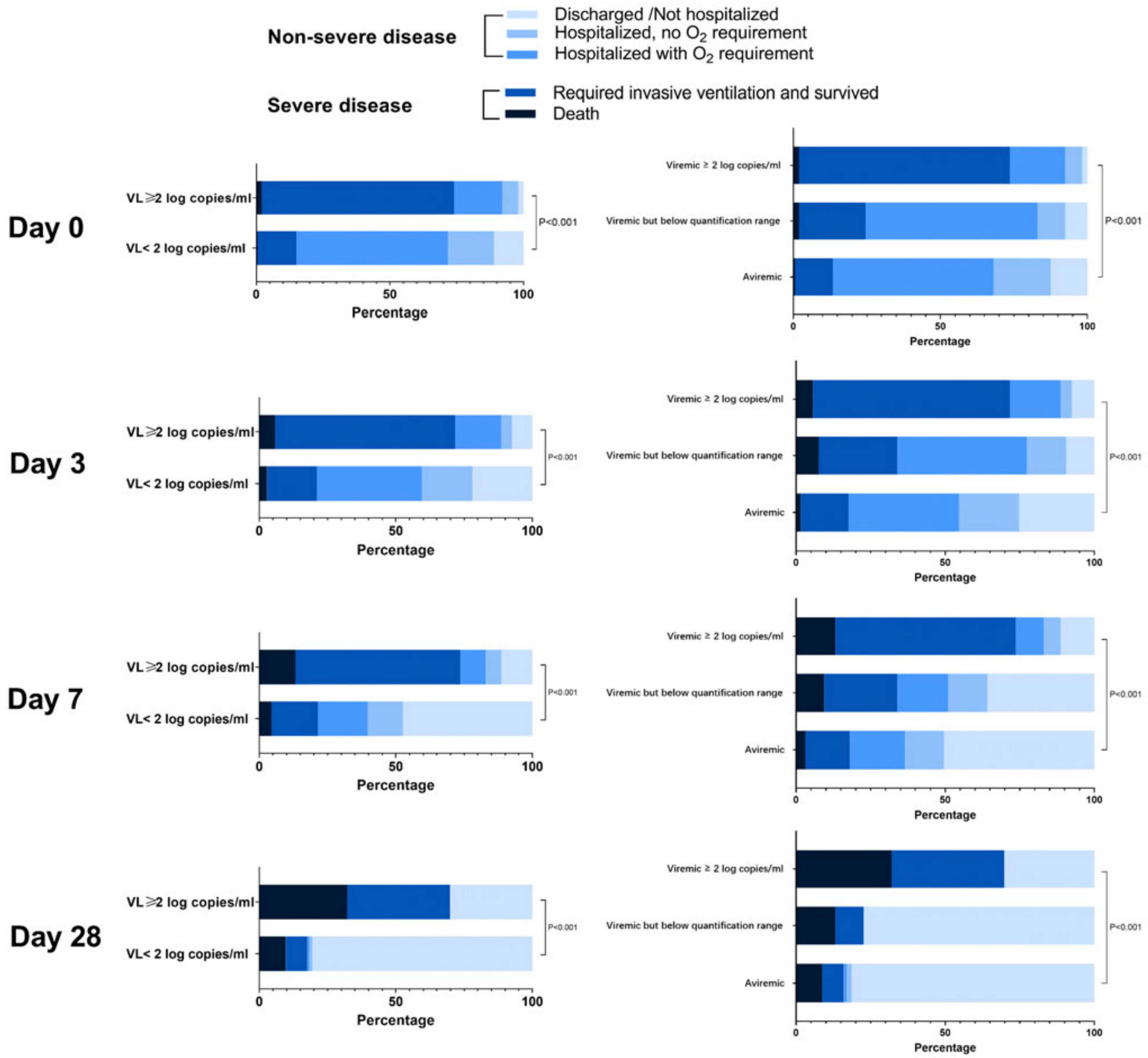

Supplementary Figure S3. Association of viremia and disease severity stratified by age groups.
P values were calculated by either chi squared test or Fisher's exact test.

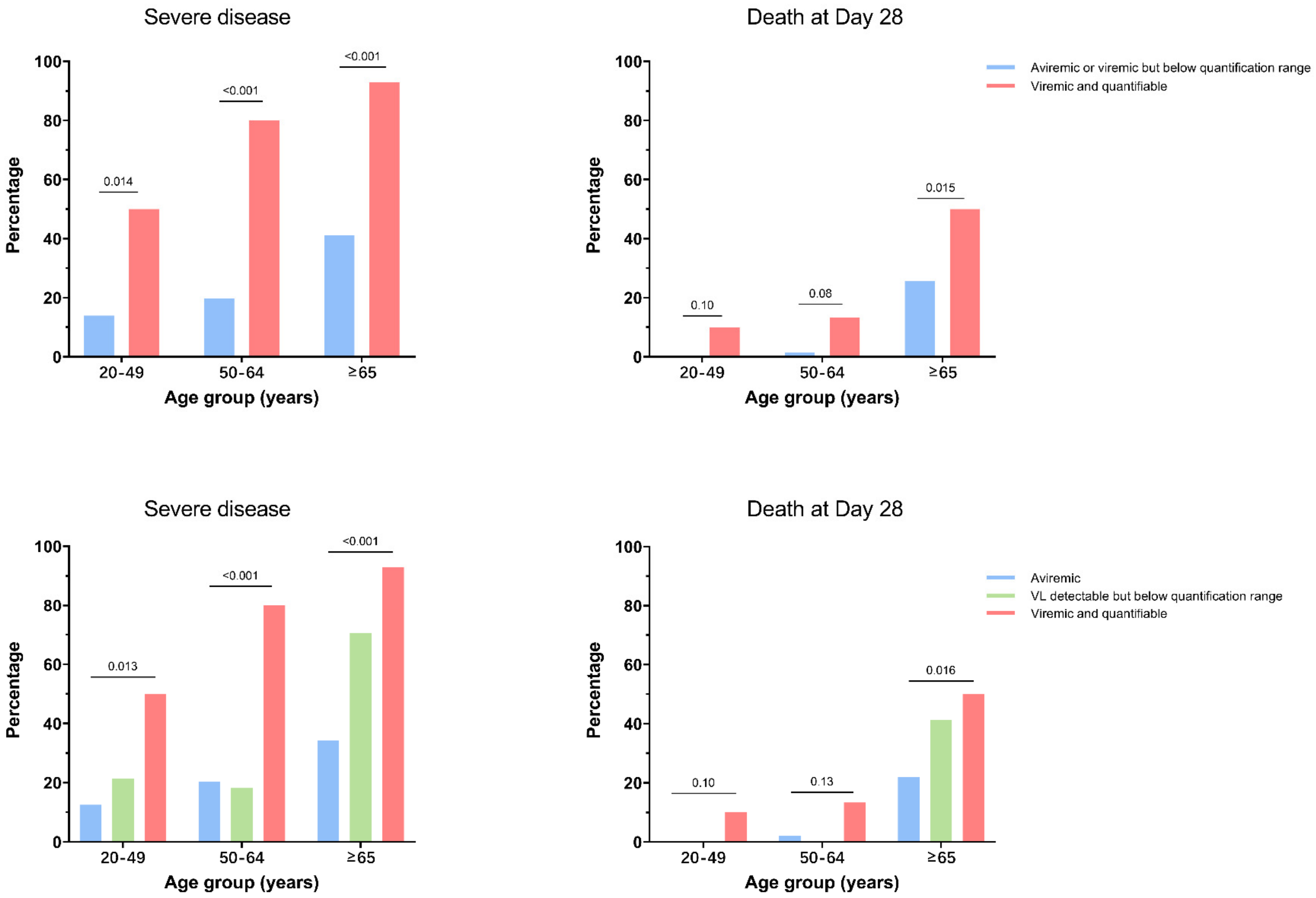

Supplementary Figure S4. 28-day mortality among different viremic groups.

Adjusted hazard ratio (aHR) was calculated using Cox Proportional regression adjusting for baseline characteristics including age, sex, race, BMI and laboratory values. Aviremic group served as reference.

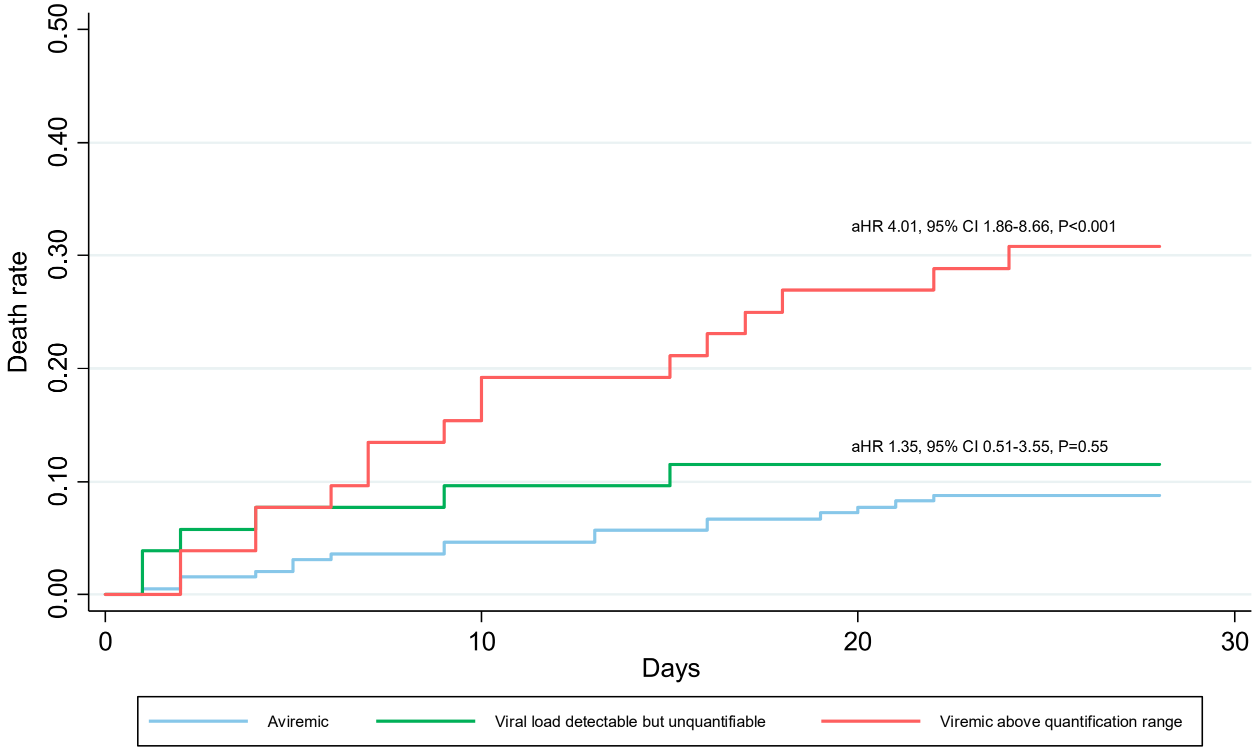

Supplementary Figure S5. Receiver operating characteristic (ROC) curve showing predictive performance of an elastic net logistic regression classifier of disease severity for Olink proteins and viremia.
Only participants with viremia above quantification range and undetectable were included in this analysis. Performance was evaluated using 100 repeats of 5-fold cross validation. Mean area under the curve (AUC) with 95% confidence intervals was 0.83, 95%CI (0.80, 0.86). Top ten predictors are listed here.

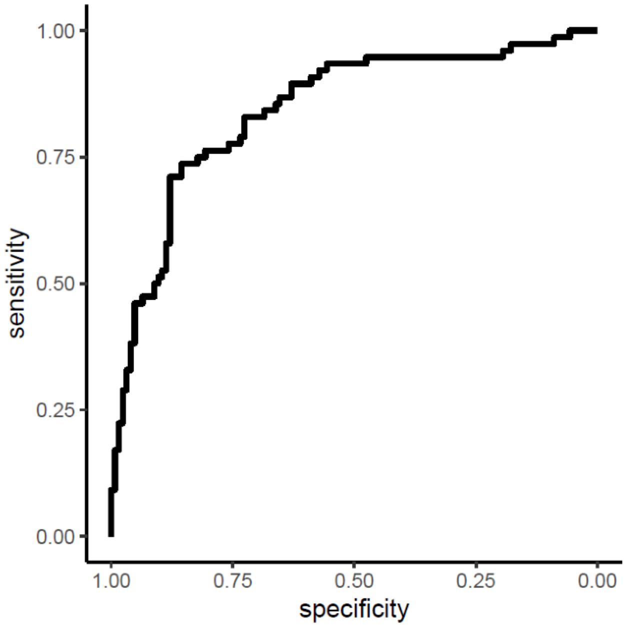

| Assay | **Mean Coefficient** | **UniProt** |
|---|---|---|
| **Viremia** | 88.0 | |
| **SEMA4D** | 39.8 | Q92854 |
| **ADGRG1** | 21.0 | Q9Y653 |
| **NDRG1** | 19.6 | Q92597 |
| **GBP2** | 10.4 | P32456 |
| **DNER** | 10.1 | Q8NFT8 |
| **TNFRSF11B** | 8.1 | O00300 |
| **KRT19** | 7.9 | P08727 |
| **FASLG** | 7.8 | P48023 |
| **IL6** | 7.7 | P05231 |

Supplementary Figure S6. Heatmap highlighting differentially expressed proteins between viremic and aviremic participants.

(A) Heatmap derived from Olink proteomics from all three time points. (B)-(D) Heatmap from Day 0, 3, and 7, respectively.

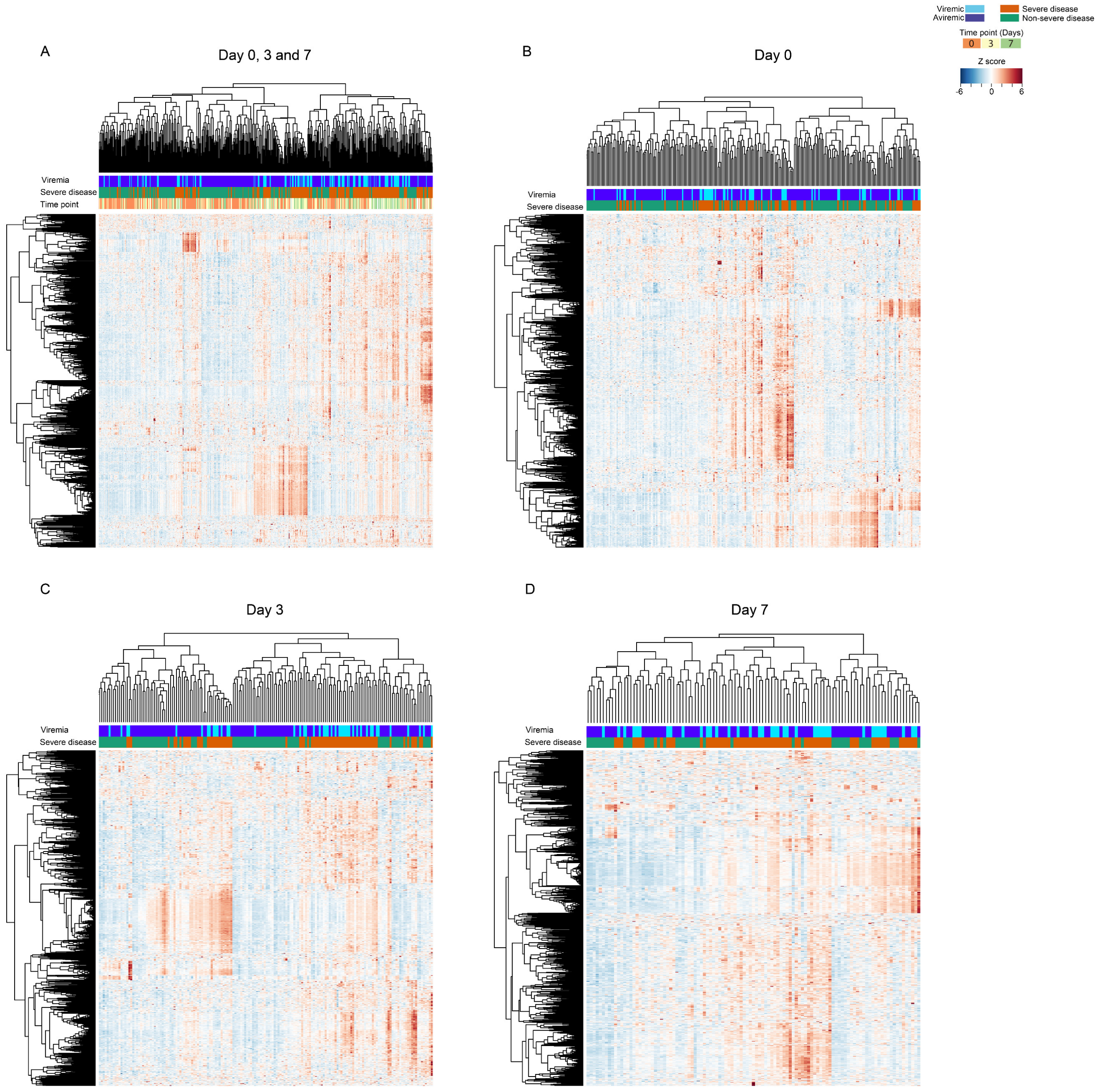

Supplementary Figure S7. Correlation between LDH, tissue-enriched protein levels, fibrosis markers, IL6, entry factors and viral load.

Spearman correlation (rho) with P value <0.05 was demonstrated in this figure.

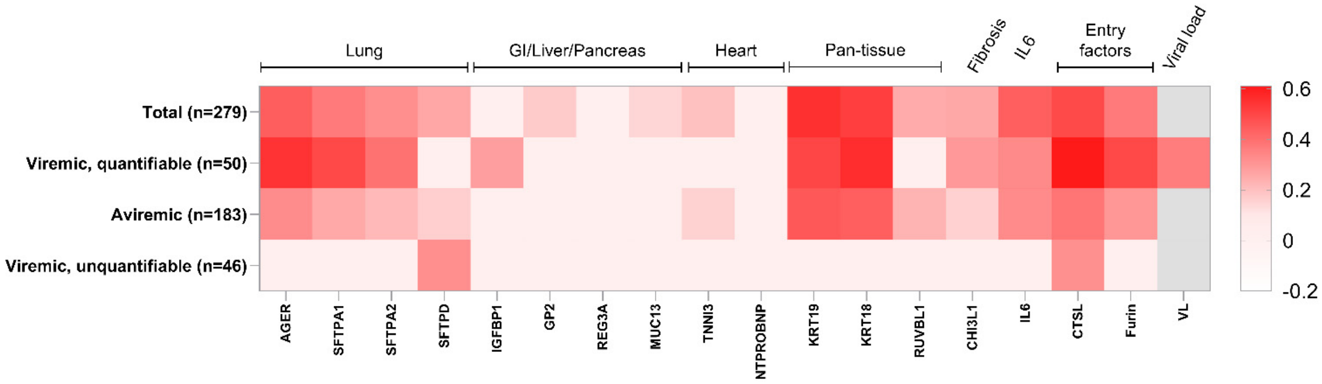

Supplementary Figure S8. Correlation between apoptosis-related protein, lung/GI tract related protein, pyroptosis-related protein and IL6.

Spearman correlation was performed in this analysis and only correlation value (rho) with a P<0.05 were shown in this graph in color (rho with P>0.05 was left in blank).

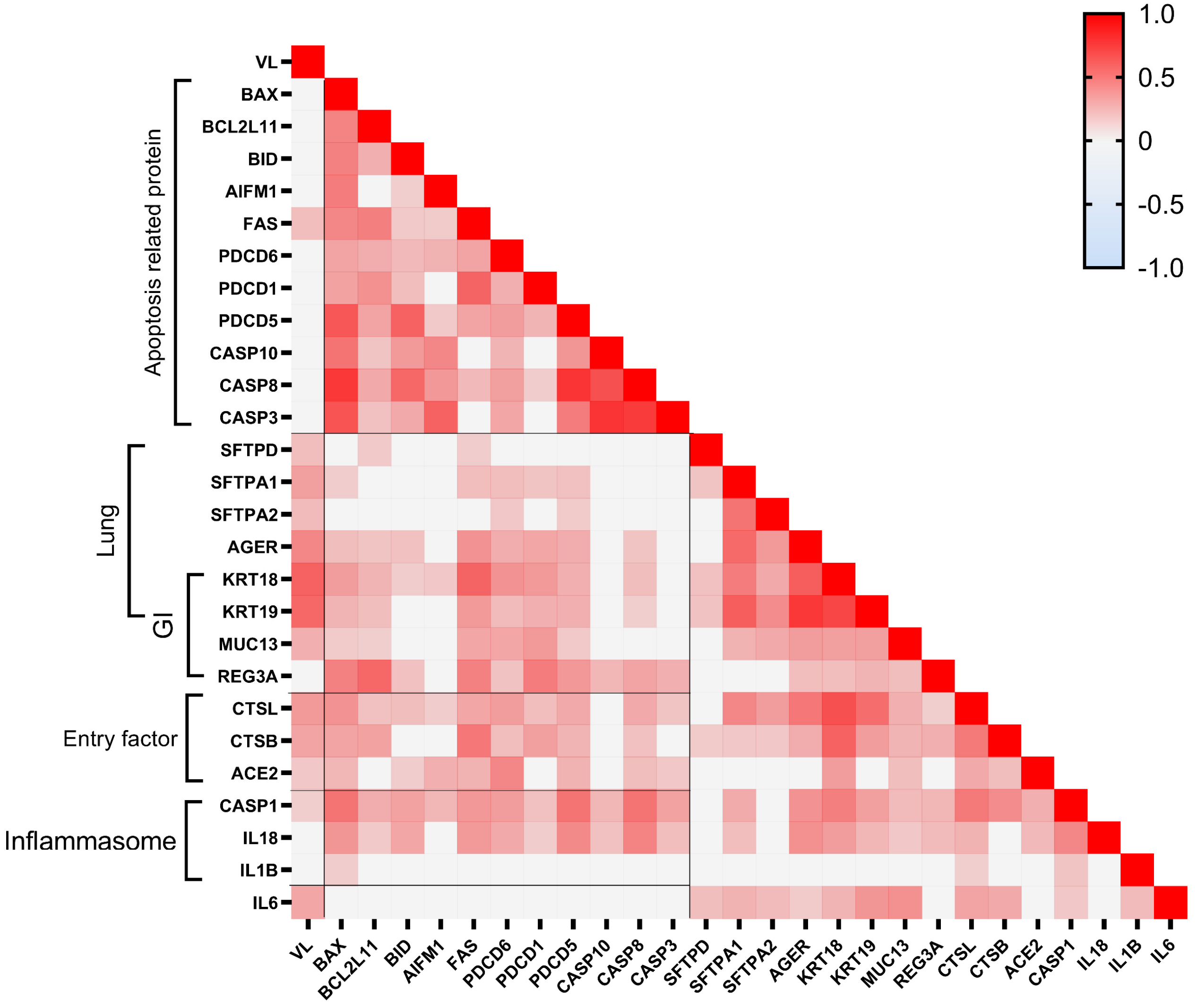

Supplementary Figure S9. Neutralization level at Day 3 and Day 7.

A subgroup of participants with undetectable viral load or detectable viral load (quantifiable or unquantifiable) at Day 3 (n=49) or Day 7 (n=39) were included in this analysis. Mann-Whitney test was used to compare the difference between two different groups. ns, not significant.

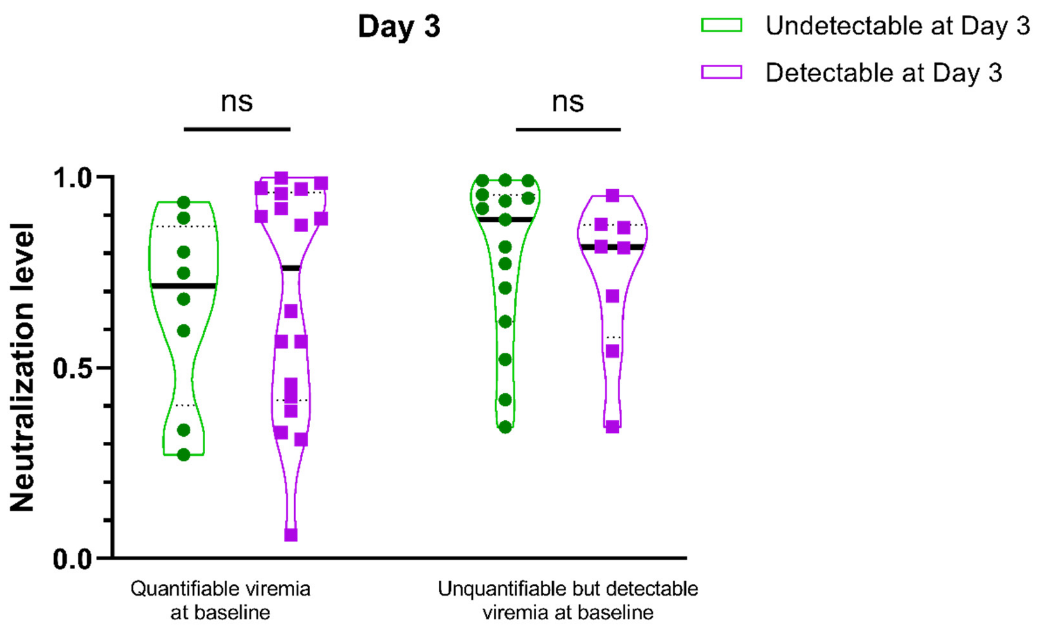

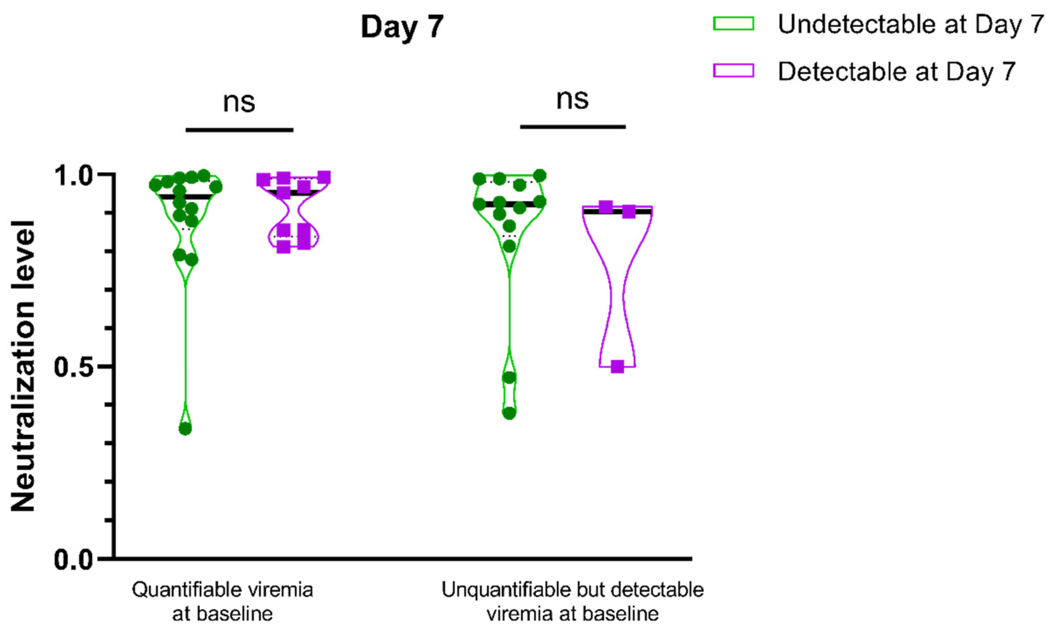
